# Turning green: the impact of changing to more eco-friendly respiratory healthcare. A carbon and cost analysis of Dutch prescription data

**DOI:** 10.1101/2021.11.20.21266571

**Authors:** Pieter ten Have, Peter Th. W. van Hal, Iris Wichers, Johan Kooistra, Paul Hagedoorn, Evelyn Brakema, Niels Chavannes, Pauline de Heer, Hans C Ossebaard

## Abstract

**Objectives:** Dry powder inhalers (DPIs) and soft mist inhalers have a substantially lower global warming potential than pressurized metered-dose inhalers (pMDIs). To help mitigate climate change, we assessed the potential emission reduction in CO2-equivalents when replacing pMDIs by non-propellant inhalers in Dutch respiratory healthcare, and estimated the associated costs.

**Design:** We performed a four-step analysis based on data from two national databases of two independent governmental bodies (the Dutch National Healthcare Institute and the Dutch Healthcare Authority). First, we calculated the number of patients with chronic obstructive pulmonary disease and asthma that were using inhalation medication (2020). Second, we calculated the number and total of daily defined doses of pMDIs and non-propellant inhalers (NPIs) that include dry powder inhalers and soft mist inhalers, as well as the number of spacers per patient dispensed by non-hospital based pharmacies in 2020. Third, we estimated the potential reduction in greenhouse gas emission if 70% of patients would switch from using pMDIs to using NPIs as eco-friendly alternatives. Fourth, we performed a budget impact analysis.

**Results:** In 2020, 1.4 million patients used inhalers for COPD or asthma treatment. A total of 460 million defined daily doses (DDDs) from inhalers were dispensed, of which – after the exclusion of nebulizers – 50.4% through pMDIs. We estimated that this usage could be reduced by 70% which would lead to an annual reduction in greenhouse gas emission of 77 - 84 million kg. CO2-eq. saving at best EUR 49.8 million per year.

**Conclusions:** In the Netherlands, substitution of pMDIs to NPIs for eligible patients is theoretically safe and in accordance with medical guidelines, while reducing greenhouse gas emission by 80 million kg. CO2-eq. on average and saving at best EUR 49.8 million per year. This study confirms the potential climate and economic benefit of delivering a more eco-friendly respiratory care.

**Strengths and limitations of this study:**

- Given availability and reliability of the data, the present analysis can easily be replicated elsewhere which allows for international comparison and aggregation.
- Implementation challenges remain underexposed.

## INTRODUCTION

Climate change is the greatest global health threat of our times, inflicting a range of ill health outcomes including (re-)emerging zoonoses such as Covid-19, non-communicable diseases and mental health disorders.[1, 2] Paradoxically, the health care industry contributes substantially to global warming. If global health care were a country, it would rank fifth for greenhouse gas emissions and its environmental footprint is substantial.[3, 4] In the Netherlands, the healthcare sector is responsible for 6-7% of the total national CO2-eq. emission.[5] Hence, the Dutch healthcare sector could play a significant role in meeting the national climate policy goals, thereby preserving planetary health and human health that depends on it.

Among the impactful solutions to deliver sustainable healthcare is the choice of inhaler type to deliver medication to the lungs of patients with asthma, allergies, or chronic obstructive pulmonary disease (COPD). Pressurized metered-dose inhalers (pMDIs) contain propellants known as hydrofluorocarbons (HFCs), potent F-gases that account for 15 megaton CO2-eq. (0.03%) of all greenhouse gas emissions worldwide (RIVM, 2021). In the European Union HFCs will be phased out by two-thirds in 2030 through limiting sale and use of air conditioning and refrigeration equipment. However, their application in metered-dose inhalers is exempted from this regulation.[6] pMDIs contain either the propellant HFC-134a or HFC-227ea. Other commonly used inhalers are dry-powder inhalers (DPIs) and soft mist inhalers. For the purpose of this paper we label these as non-propellant inhalers (NPIs). These are as safe and effective in most patients but do not contain greenhouse gases which is why the life cycle assessment of their environmental impact is substantially lower than those of pMDIs.[7]

### BOX 1.

**Global Warming Potential (GWP)**

The global warming potential is the heat absorbed by any greenhouse gas in the atmosphere compared to the mass of CO_2_. The GWP of CO_2_ is 1.0. The GWPs of HFC-134a and HFC-227ea, hydrofluorocarbons used in metered-dose inhalers, are 1,330 and 3,220.

Several studies have assessed the costs and benefits of switching to medication with a lower global warming potential (see Box 1). Wilkinson et al. found considerable reductions in both CO2 emissions and pharmaceutical costs.[8] Janson et al. recommend that “the lower carbon footprint of DPIs should be considered alongside other factors when choosing inhaler devices.”[9] In their review, Starup-Hansen et al. recommend to update guidelines: “guidance should consider the potential benefits of advising DPIs as the device of choice in new diagnoses of asthma and COPD as well as the benefits of switching patients currently using pMDIs to DPIs where clinically appropriate.”[10] These recommendations have been recently adopted in the guidelines ‘Asthma in adults’[11] and ‘COPD’[12] of the Dutch College of General Practitioners. Among other updates, these guidelines contain the same modest, though historical, reference to considering the environmental impact of the medicine of choice for the prescribing physician (see Box 2).

### BOX 2.

**NHG-Guidelines ‘Asthma in adults’ (2020) and ‘COPD’ (2021)**

#### One of the criteria in de decision aide for choosing an inhaler device

*“A general objection against metered-dose inhalers is that they contain a greenhouse gas with a strong environmental impact*.*”*

#### Note

*“Metered-dose inhalers use HFC propellants. The F-gas hydrofluorcarbon does not affect the ozone layer but is a strong greenhouse gas. The environmental impact of 1 inhalation is 25 times larger than a dry-powder inhalation. Environmental impact of production, transport and waste processing (*..*) have not been included*.*”*

To understand the implications of changing from pMDI to more eco-friendly NPI-use for policy, practice, and patients in settings, we build on the cost and carbon analysis of Wilkinson et al.[8]. In this paper, we calculated the environmental impact of this change in Dutch primary and secondary respiratory healthcare and analyzed the associated pharmaceutical and device costs.

## METHODS

We performed a four-step data analysis of prescription data in order to estimate the carbon equivalent footprint of prescribed inhalers over a one-year period (2020). We determined how much inhalation medication could be attributed to the following patient groups: 1) asthma, 2) COPD, 3) severe COPD and 4) children younger than 7 years of age. Estimations were based on the GIP database (Genees-en hulpmiddelen Informatie Project | Medicines and medical devices Information Project) of the Dutch National Health Care Institute and the DIS database (DBC Informatie Systeem | Diagnosis-Treatment Combination Information system) of the Dutch Healthcare Authority, both independent government bodies residing under the Dutch Ministry of Health, Welfare and Sports. GIP is a representative information system containing data on the use and cost of prescription drugs and medical devices.[13] DIS contains information of all treatment trajectories in Dutch medical specialist care, including pulmonary medicine, mental health care, forensic care and rehabilitation.[14] Health care providers are legally required to deliver these data for policy making and regulation. A *Supplementary File* contains the complete data analysis protocol and additional information regarding methodological details, assumptions and choices made.

First, we calculated the number of patients with asthma or COPD that used inhalation medication in the Netherlands in 2020 by joining diagnoses codes to inhalation medication. Second, we calculated the number of defined daily doses (DDDs) discriminating between pMDIs and NPIs. Nebulizers were excluded from the analysis since they do not contain propellants and due to their size and dependency on electricity, they are not to be considered an alternative to pMDIs for use by patients at home. We included the soft mist inhalers in de NPI group, because they do not contain propellants and may be considered an alternative to pMDIs. Third, we determined the volume of pMDIs that could hypothetically be replaced by NPIs in a safe and medically responsible way. We estimated the size of this volume in DDDs, according to current medical guidelines excluding children younger than 7 years of age and those patients with severe COPD having at least two exacerbations per year. In our data the subgroups ‘younger than 7 years’ and ‘severe COPD’ consume 14.3% of the total medication delivered by pMDI. Hence, if we would disregard their pMDI-use, and only replace inhalers of the remaining patients, we could theoretically achieve a 85.7% reduction of pMDI-use. In these two subgroups (younger than 7, severe COPD), it is possible to safely replace pMDIs in inhalation corticosteroid (ICS) maintenance therapy for NPIs, without any negative medical impact. Here, breathing is not hampered during maintenance therapy and an immediate effect of ICS is not required. We nonetheless choose a more conservative estimate of change. We used the frequently stated figure of 10% pMDI-use in Sweden as a proxy, assuming Sweden and The Netherlands are comparable in terms of a variety of social-epidemiological indicators.[15, 16] Hence it is likely that the latter country could approach Sweden’s level of NPI-prescription to an again more conservative, putative 15%. From the current level of 50.4% down to 15% pMDI-use equals a 70% reduction, which is considerably less than the previous 85.7%. Based on our data we know how many canisters of each type were prescribed in 2020, and we applied two conversion tables, one published by Wilkinson et al.[8] and the other one by Jeswani & Azapagic.[7] Since they use different resources for quantification we have used a range instead of an average. Finally we calculated the kg. CO2-eq. decrease as a consequence of this substantial 70% reduction in pMDI-use. In the fourth and last step we calculated if this potential replacement could be achieved in a cost-neutral way. By determining both the current costs of medication, spacers and estimated replacement costs we calculated the difference. For the replacement costs we applied two realistic scenarios, one is the low-cost scenario in which pMDIs are replaced by low-cost NPIs. In the second scenario pMDIs are replaced by average-cost NPIs. People living with COPD or asthma were not involved in the design and conduct of this study.

## RESULTS

In 2020, 1,392,743 patients used inhalation medication in the Netherlands, and they received a total of 459,621,151 defined daily doses (Table 1). In addition 549,224 spacers were administered to 509,650 (pMDI-using) patients meaning that 61% of 831,494 pMDI-using patients could use their inhaler together with a yearly to-be-replaced spacer, as recommended by Dutch medical guidelines.

**Table 1.** Inhalation medication in the Netherlands 2020

| *Inhaler type* | *Number of patients \** | *Number of DDDs \*\** | *% of total DDD use* |
| --- | --- | --- | --- |
| pressurized metered-dose (pMDI) | 831,494 | 228,074,487 | 50.4% |
| non-propellant (NPI) | 798,598 | 224,615,426 | 49.6% |
| nebulizers (excluded in further analysis) | 23,479 | 6,931,238 |  |
| pMDI and/or NPI (included) | 1,388,255 | 452,689,914 | 100% |
| pMDI and/or NPI and/or nebulizers (total group) | 1,392,743 | 459,621,151 |  |

\* Users may use different types of inhalers at the same time
\*\* Defined daily dose

After excluding the use of nebulizers, we focused on the group of 1,388,255 patients using pMDI and/or NPI, who were prescribed over 452,689,914 DDDs in 2020 (Table 1).

The total amount of medication delivered in 2020 by pMDI is 228,074,487 DDDs. We observed that 50.4% of the medication has been delivered using pMDIs, 49.6% per NPIs (Table 1).

Not all inhalation medication is delivered by both types of inhalers and can be switched. Long-acting muscarinic-antagonists (LAMA) and the combination of long-acting beta agonists with long-acting muscarinic-antagonists (LABA-LAMA) were only available as NPI, the combination of short-acting beta agonists with short-acting muscarinic-antagonists (SABA-SAMA) was only available as pMDI.

The number of patients that could hypothetically switch safely to NPIs with the same content would be using 154,647,286 DDDs, equal to 4,500,234 canisters.

Using the Wilkinson’s conversion table with “mg HFC per canister”, delivers a reduction of 84 million kg. CO2-eq.[8] Using the conversion table from Jeswani & Azapagic yields a reduction of 77 million kg. CO2-eq.[7] The range being 76,959,654 – 83,817,348 kg. CO2-eq. with an average of 80,388,501 kg. CO2-eq. corresponding to 61,101 kg. HFC; HFC-134a for the better part (Figure 1).

**Fig. 1.**
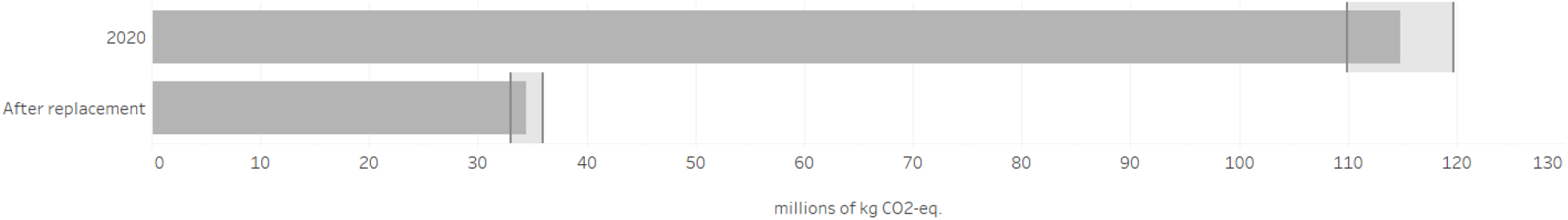
Environmental impact (in kg. CO2-equivalents) of a hypothetical replacement of pMDIs in The Netherlands.

We calculated if shifting to NPIs could be achieved in a cost-neutral way. We determined both the current costs of medication and spacers, we estimated replacement costs and we calculated the difference. For the replacement costs we applied two realistic scenarios. One is a low-cost scenario in which pMDIs are replaced by low-cost NPIs. In the second scenario pMDIs are replaced by average-cost NPIs in current market share (Table 2).

**Table 2.** DDD Volumes, costs of medication and spacers

|  | pMDI-use in 2020, in medication groups: SABA, LABA, ICS, SAMA, LABA-ICS, LABA-SAMA-ICS* | 70% of pMDI-use (part that can theoretically be safely replaced) | Replacement of pMDI by low-cost NPI | Replacement by NPI, in current market share |
| --- | --- | --- | --- | --- |
| Volume in DDD | 220,924,694 | 154,647,286 | 154,647,286 | 154,647,286 |
| Medication costs | € 171,372,329 | € 119,960,630 | € 82,861,418 | € 141,025,299 |
| Costs of spacers | € 18,095,091 | € 12,666,564 | n.a. | n.a. |
| Total costs | € 189,467,420 | € 132,627,194 | € 82,861,418 | € 141,025,299 |

\*
SABA = short-acting beta agonists | SAMA = short-acting muscarinic-antagonists LABA = long-acting muscarinic-antagonists | LAMA = long-acting muscarinic antagonists ICS = inhalation corticosteroids

If the percentage of DDD’s from pMDI could be reduced from 50.4% to 15% this 70% reduction implies a decrease of 154,647,286 DDDs which equals EUR 132,627,194 (medication + inhalers cost EUR 119,960,630 plus the cost of spacers EUR 12,666,564). Replacing this by low-cost NPIs, would incur a cost of EUR 82,861,418 saving approximately EUR 49.8 million annually. The average-cost scenario would result in EUR 8,4 million annual added expenses.

## DISCUSSION

The healthcare sector needs to decrease greenhouse gas emissions to help mitigate climate change. This may be viewed as a moral and practical obligation in times of climate crisis and the global health emergency it implies.[17] To achieve this, substantiated and medically safe eco-friendly alternatives are necessary. In this study, we assessed the hypothetical impact of converting eligible patients from using pMDIs to using NPIs in the Netherlands, both in terms of greenhouse gas emissions and in cost. With these outcomes we seek to offer insight into the impact of making this change and to inspire health care professionals to act climate responsibly which is congruent with announcements of professional organisations such as the British Thoracic Society,[18] the European Respiratory Society,[19] the International Society for Quality in Health Care,[20] and the US National Academy of Sciences, Engineering, and Medicine.[21]

Our results show that a sizeable reduction in greenhouse gas emissions is attainable in the Netherlands with a readily available eco-friendly alternative. The financial impact of this shift depends on the choice for either a low-cost option or a more expensive option, but we demonstrated a cost reduction is feasible. The estimated cost-saving does not include financial calculations of patient training or potential drawbacks of substitution such as lower adherence leading to increased GP visits or hospital admissions.

These results are in accordance with earlier studies [8, 9, 22] but we were relatively stringent in our eligibility criteria (which patients are able to change safely) and more selective as to what brands to include for the financial impact estimation. Obviously the outcomes refer to Dutch respiratory health care, its specific patient population and medication use.

In estimating the environmental impact of pMDIs, we considered their full amount of propellants. We did not subtract unknown quantities of propellants that may remain in the canister after use, assuming that sooner or later 100% of these gases will be released into the atmosphere. We did not include other environmental impacts of pMDIs nor NPIs, as would have been done in a full life cycle assessment (LCA). LCAs typically include the whole spectrum of production, packaging, distribution, distribution, usage, waste, etc. However, pMDIs’ global warming effect is mainly caused by their use (95-98%), not by the manufacturing of this class of inhalers.[7, 8] Though NPIs, as opposed to pMDIs, generate much lower GWP, LCAs imply other harmful impacts that eventually should be included in a comparison such as human toxicity, marine eutrophication or fossil depletion.[7]

Our study implies that if medically safe and possible, choosing the medicine or device with the least environmental impact is imperative in times of global climate crisis. This is not just about patients’ choice, as may be suggested by NICE’s patient decision aid.[23] It could be considered the prescriber’s task as well. Therefore it should be integrated in medical guidelines and standards as part of health care quality improvement trajectories much like Mortimer et al. have elegantly proposed and practiced.[24] This should not affect the established fact that suitable patient training and monitoring of inhalation techniques are a sine qua non for effective inhaler use for all a patients, especially for children.[25, 26] In the Netherlands, general practitioners recently updated their guidelines on the management of asthma and COPD, and included a recommendation to consider the environmental impact of the medicine of choice (see Textbox 2). In view of the health emergency represented by the climate crisis we recommend that pulmonologists also consider to update national and local guidelines and appreciate the potential benefits of advising green inhalers as the device of choice in new diagnostics of asthma and COPD and the benefits of resetting patients currently using pMDIs to NPIs if safe and possible. In 2019 Belgian pulmonologists recommended the use of DPIs to lung patients not just because they can deliver better treatment results for asthma and COPD but also because they are “far less damaging to the environment than traditional propellant driven aerosols.”[27]

Evidently, the chosen medication should be fitting for the individual patient. It is beyond the scope of this study to include all specific circumstances in which patients cannot use NPIs. Since daily use and emergency use are quite different, there have been reservations about DPIs in case of exacerbations especially since both the expiratory flow and the inspiratory (‘trapped air’) flow of breath are obstructed leading to patients’ preference for pMDIs in such circumstances. In Sweden soft mist inhalers are recently used more often in such cases because they require minimal inspiratory power. Wilkinson et al. referring to a data analysis of the NHS Business Services Authority, suggest that in England “clinicians believe the vast majority of patients can use a DPI effectively.”[8]

Apart from climate and economic benefits we identified more advantages of replacing pMDIs with NPIs as suggested by research and practice (Table 3).

**Table 3.** Plausible advantages of replacing pMDIs with DPIs.

| Plausible advantages | References (if present) |
| --- | --- |
| Less critical errors are made using DPIs as compared to pMDIs. | Chrystyn H, van der Palen J, Sharma R, Barnes N, Delafont B, Mahajan A, Thomas M. Device errors in asthma and COPD: systematic literature review and meta-analysis. *NPJ Prim Care Respir Med*. 2017 Apr 3;27(1):22. |
| Sometimes pMDIs are used when empty, which may lead to poor disease control and less quality of life. | Conner JB, Buck PO. Improving asthma management: the case for mandatory inclusion of dose counters on all rescue bronchodilators. *J Asthma*. 2013 Aug;50(6):658-63. doi: 10.3109/02770903.2013.789056. Epub 2013 Apr 29.<br>Tsangarides A, Wilkinson A, Mir F. Disadvantages of salbutamol pressurised metered-dose inhalers (pMDIs). *Thorax* 2018;73:A193-A194. |
| Some pMDIs are unknowingly considered empty and are disposed of leading to unnecessary costs. | Holt S, Holt A, Weatherall M, Masoli M, Beasley R. Metered-dose inhalers: a need for dose counters. *Respirology (Carlton, Vic.)*. 2005 Jan;10(1):105-106. |
| Following Dutch clinical guidelines, pMDI-users should yearly receive a new spacer. During 2020 however, only 61% of pMDI-using patients received it which implies suboptimal quality of care. |  |
| Changing to DPI may improve guideline adherence because use of a spacer is not required for DPI. |  |
| Use of DPI requires no spacers and consequently does at least not generate nonreusable plastics |  |

The present study does not discuss implementation questions, or probable (dis-)advantages of both pMDIs and NPIs. It is certainly useful to address the preferences and prejudices of patients and professionals and we know that citizens, patients and professionals are increasingly willing to choose eco-friendly alternatives but there is no knowledge on this specific shift from pMDIs to NPIs.[28-30] Next to that, while some practical (dis-)advantages of both pMDIs and NPIs are known we recommend explaining these to patients similar to the NICE decision aid as well as to professionals.[23, 31] For example, most pMDIs do not have dose-counters. While all DPIs have a counter they do not necessarily prevent using an empty device. Without a dose-counter it may be hard to know how many doses are left in the device. Unknowingly using empty pMDIs could lead to avoidable exacerbations or even avoidable hospital admissions. Unknowingly replacing pMDIs that still contain medication would incur unnecessary cost.[32] Adherence to inhalation instructions may be an issue when it comes to changing, but this is already an issue e.g., not every patient with an pMDI uses the recommended, though bulky, spacer. Also, adherence to inhalation medication therapy should be promoted by repeated inhalation instruction.[33] Switching without sufficient instruction may result in uncontrolled, exacerbations and increased use of health care services. Uniformity of the devices in case of multiple inhaler use is relevant here. Such questions pertain to responsible implementation, a subject we will address in our follow-up study in the context of Covid-19 recovery plans.

The pharmaceutical industry meanwhile continues to develop and study inhalers with lower climate impacts. And new propellants will enter the market. For patients who are dependent on pMDIs, this is meaningful. Given that these developments have not yet entered the market and knowledge of these is still limited, we will not elaborate on this matter. Research should nonetheless include more green metrics into their output and outcome parameters. This would enable meta-analyses and evidence-based climate-responsible innovation in health care.

## CONCLUSIONS

Large scale replacement of pMDIs with NPIs would have a substantial climate impact in respiratory healthcare. In 2020 about 1.4 million patients using pMDI and/or NPI, were prescribed over 460 million DDDs. The use of pMDIs is more or less equally prevalent among patients with COPD and patients with asthma. Half (50.4%) of the medication has been delivered through pMDIs that have a relatively high global warming potential. The percentage of NPI-delivered inhalation medication that can safely be replaced is estimated to be 70%, resulting in an environmental health benefit of 80,388,501 kg. CO2-eq. on average, which equals the carbon dioxide emission of just over 10718 Dutch households. This shift could be achieved with low budgetary risk. In the low-cost scenario it may even lead to a cost reduction of approximately EUR 49.8 million per year in Dutch respiratory health care. The average-cost scenario would result in EUR 8,4 million annual added costs while still reducing greenhouse gas emission.

## Supporting information

SupplFile Data analysis protocol

## Data Availability

All data produced in the present work are contained in the manuscript or in its attachments

## Twitter

@hcointox

## Acknowledgements

The authors would like to thank the National Health Care Institute for their support in drafting this paper.

## Contributors

All authors meet the required criteria for authorship. PtH contributed to the conception, designed the study, did most of the analysis and interpretation of the data. PvH, IW, JK, PdH, NC, EB, PH helped analyse and interpret the data for health care practice and implementation issues, and revised the manuscript. HCO contributed to the conception, drafted the manuscript and revised it for intellectual content.

## Patient consent for publication

Not required.

## Provenance and peer review

Not commissioned; to be externally peer reviewed.

## Data availability statement

Both databases used for this article are publicly accessible. Commercially sensitive information related to brand names cannot be made available.

## Author note

All authors meet the required criteria for authorship: substantial contributions to the conception or design of the work; the acquisition, analysis or interpretation of data for the work; drafting the work or revising it critically for important intellectual content; final approval of the version to be published and agreement to be accountable for all aspects of the work in ensuring that questions related to the accuracy or integrity of any part of the work are appropriately investigated and resolved.

## Supplementary File

Data analysis protocol (Appendix)

