## Supplementary material for "Turning green: the impact of changing to more eco-friendly respiratory healthcare. A carbon and cost analysis of Dutch prescription data": SupplFile Data analysis protocol

Ten Have P, Van Hal P, Wichers I, Kooistra, J, Hagedoorn P, Brakema EA, Chavannes NH, De Heer, P & Ossebaard HC (2021) Turning green: the impact of changing to more eco-friendly respiratory healthcare. A carbon and cost analysis of Dutch prescription data. Corresponding author: Hans C Ossebaard PhD, National Health Care Institute, P.O. Box 320, 1110 AH, Diemen, The Netherlands.

### SUPPLEMENTARY FILE: DATA ANALYSIS PROTOCOL

*Introduction*

We calculate the impact of replacing pressurized metered-dose inhalers (pMDIs) by non-propellant inhalers (NPIs), a group consisting of both dry powder inhalers (DPIs) and soft mist inhalers, on greenhouse gas emissions in Dutch respiratory healthcare. The major steps of our method are shown in Figure 1.

Figure 1. Steps to calculate the impact of conversion of pMDI to NPI

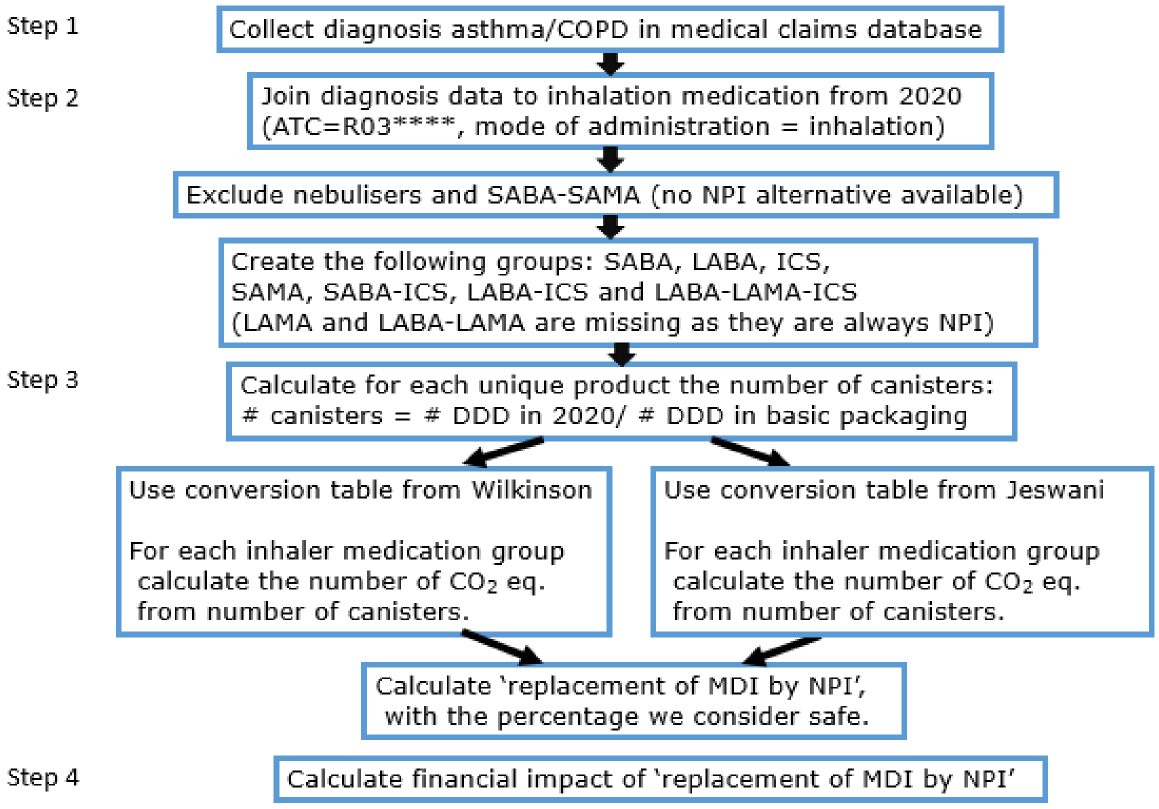


Our data-analysis protocol is:

**Step 1: Collect diagnoses asthma/COPD from medical claims database**

Use the DIS database (DBC Informatie Systeem | Diagnosis-Treatment Combination Information system) to collect the identifiers and diagnoses of patients that received care for asthma or COPD between 2012 and 2020. The DIS database is a medical claims database covering all medical care delivered to Dutch citizens, including private health care. The independent government body Dutch Health Care Authority (Nederlandse Zorgautoriteit) owns this database. The reason for this initial step is to find out how many pMDI DDDs were prescribed for patients with severe COPD, as guidelines do not consider them eligible for DPI. Also, we wanted to know how pMDIs are distributed between asthma and COPD. The DIS database does not contain all primary care diagnoses.

**Step 2: Join these diagnoses to the inhalation medication prescribed in 2020**

The GIP database (Genees- en hulpmiddelen Informatie Project | Medicines and medical devices Information Project) contains all prescriptions of all Dutch citizens from pharmacies since about 1985. The Dutch National Health Care Institute (Zorginstituut Nederland) is the owner of this database. Use the GIP database to select all medication where the ATC-code starts with 'R03', the mode of administration is 'inhalation' and the year is 2020. Increase all numbers with 3%, because a few small health insurance companies do not deliver claims data. These missing data represent 3% of the claims volume.

Exclude the nebulizers since they don't contain propellants and because they are usually not an appropriate alternative for a pMDI due to their size and dependency on electrical energy.

Label 'soft mist inhalers' and 'DPIs' as non-propellant inhalers (NPIs) since they do not contain propellants and may be considered an alternative to pMDI.

Exclude the SABA-SAMA medication, because there are no NPIs containing both SABA and SAMA and they can't be replaced properly. We considered all replacements from pMDI to NPI to be acceptable as long as the medication group stays the same and the patient doesn't end up with more inhalers. Because there is no NPI SABA-SAMA available, replacing a pMDI SABA-SAMA by a NPI SABA plus a NPI SAMA, would lead to an extra inhaler. This, we did not consider acceptable for replacement. We believed it is not necessary to keep the ATC-code the same during a replacement. E.g., we considered replacing any pMDI SABA by any NPI SABA to be acceptable because the medication group did not change.

Create the following medication groups, allowing replacement within each group: SABA, LABA, ICS, SAMA, SABA-ICS, LABA-ICS and LABA-LAMA-ICS. LAMA and LABA-LAMA are missing from the list of inhalation medication with propellants, as they are always delivered by NPI.

**Step 3: Calculate the carbon dioxide impact of replacement of pMDI by NPI**

Calculate the number of canisters for each specific inhalation medication product. The number of DDDs in basic packaging is one of the database fields of the GIP database.

Calculate the carbon dioxide equivalent ($CO_2$ eq.) per type of canister. Do this once by using the conversion table from Wilkinson et al.[1] and once by using the conversion table from Jeswani & Azapagic.[2] Because the two conversion tables deliver different results, we choose to use both tables in order to create a range. Not all types of canisters were mentioned in the two conversion tables. Therefore we added some assumptions to the tables and marked them. We based these assumptions on the other data.

Table 1. Conversion table adapted from Wilkinson et al. (2019)

| *Inhalation medication group* | *kilogram CO2 per canister* |
|---|---|
| ICS | 20.4 |
| LABA | 15.6 |
| LABA-ICS, Flutiform | 36.5 |
| LABA-ICS, all others | 19.6 |
| LABA-LAMA-ICS (assumption) | 19.5 |
| SABA | 17.2 |
| SABA-ICS (assumption) | 19.5 |
| SAMA | 14.3 |

[1] Wilkinson AJK, Braggins R, Steinbach I, *et al*. Costs of switching to low global warming potential inhalers. An economic and carbon footprint analysis of NHS prescription data in England. *BMJ Open* 2019;9:e028763. doi:10.1136/ bmjopen-2018-028763

[2] Jeswani, H. K., & Azapagic, A. (2019). Life cycle environmental impacts of inhalers. *Journal of Cleaner Production*, *237*, [117733]. https://doi.org/10.1016/j.jclepro.2019.117733

Table 2. Conversion table adapted from Yeswani & Azapagic (2019)

| *Inhalation medication group* | *kilogram CO2 per canister* |
|---|---|
| ICS, brand = Alvesco, ATC = R03BA08 | 10.946 |
| ICS, all others | 15.0475 |
| LABA | 13.2535 |
| LABA-ICS, Flutiform | 32.0048 |
| LABA-ICS, all others | 14.508 |
| LABA-LAMA-ICS (assumption) | 19.5 |
| SABA, brand = Airomir_ | 7.696 |
| SABA, all others | 23.374 |
| SABA-ICS (assumption) | 19.5 |
| SAMA | 14.17 |

Calculate the impact of a 70% decrease of pMDI use. In 2020 in the Netherlands 50.4% of inhalation medication DDDs consist of pMDIs. We assume this can safely be lowered to 15%, which is equal to a 70% decrease ((50.4% - 15%)/50.4%). We have two arguments for this assumption:

1) Current Dutch COPD-guidelines[3] state that children younger than 7 years and patients with at least two exacerbations per year are more dependent on pMDIs. Children cannot yet coordinate their breathing well and need an pMDI and a spacer, and patients with 'severe' COPD have a low inspiratory flow and therefore require the force of a pMDI propellant. We defined 'severe COPD' as COPD requiring at least 42 DDDs of oral corticosteroids per year, which is equal to two treatments of exacerbations. In our data we observed that 14.3% of pMDI DDDs were prescribed for patients who were either younger than 7 years or had severe COPD. If we leave their pMDI DDDs untouched, a replacement of 85.7% would theoretically be possible (100 - 14.3)/100).

[3] Bischoff E, Bouma M, Broekhuizen L, Donkers J, Hallensleben C, De Jong J, Snoeck-Stroband J, In 't Veen JC, Van Vugt S, Wagenaar M. NHG | Nederlands Huisartsen Genootschap (2021) *NHG-richtlijn COPD* [Dutch College of General Practitioners Guideline COPD]. Available: https://richtlijnen.nhg.org/standaarden/COPD [Accessed 19 Apr 2021].

2) In Sweden approximately 10% of inhalation medication consists of pMDIs. [4] Assuming Sweden and The Netherlands are comparable in terms of a variety of social-epidemiological indicators we believe the latter country should be able to lower their percentage of DDDs delivered by pMDIs to 15%.

**Step 4: Calculate the financial impact or replacement from pMDI to NPI**

Calculate the financial impact with two scenario's:

1) Low-cost scenario
   Calculate the costs of all pMDI medication and spacers in 2020. Add these costs and multiply by the replacement percentage of 70. These are the current costs.
   Divide the pMDI medication into the groups: SABA, LABA, ICS, SAMA, LABA-ICS and LABA-LAMA-ICS. Within each group calculate the costs if 70% of pMDI DDDs would be replaced by the low cost NPI in the same group. These are the replacement costs.

2) Average-cost scenario
   Calculate the costs of all pMDI medication and spacers in 2020. Add these costs and multiply by the replacement percentage of 70. These are the current costs.
   Divide the pMDI medication into the groups: SABA, LABA, ICS, SAMA, LABA-ICS and LABA-LAMA-ICS. Within each group calculate the costs if 70% of pMDI DDDs would be replaced by the weighted average cost of NPI of the same group. These are the replacement costs.

[4] Lavorini F, Corrigan CJ, Barnes PJ, Dekhuijzen PR, Levy ML, Pedersen S, Roche N, Vincken W, Crompton GK; Aerosol Drug Management Improvement Team. Retail sales of inhalation devices in European countries: so much for a global policy. Respir Med. 2011 Jul;105(7):1099-103.

#### Outcome of steps 1 and 2

Table 3: Inhaler use by diagnosis (nebulisers were excluded, soft mist inhalers were included within DPI).

| *Type of inhaler* | *Patient has diagnosis* | *Number of patients/ users ** | *Number of DDDS of inhalation medication* | *pMDIs prescribed for asthma* | *pMDIs prescribed for COPD* |
|---|---|---|---|---|---|
| pMDI | n.a. | 501,058 | 81,324,887 | | |
| NPI | n.a. | 455,706 | 88,850,395 | | |
| pMDI | Asthma | 157,475 | 63,011,726 | 63,011,726 | |
| NPI | Asthma | 121,293 | 36,650,900 | | |
| pMDI | COPD | 151,786 | 72,460,301 | | 72,460,301 |
| NPI | COPD | 201,022 | 90,408,417 | | |
| pMDI | Asthma and COPD | 21,176 | 11,277,574 | 5,638,787 | 5,638,787 |
| NPI | Asthma and COPD | 20,577 | 8,705,713 | | |
| total | | | 452,689,914 | 68,650,513 | 78,099,088 |

It is clear that pMDI use is not very different between asthma and COPD. It is also clear that primary care diagnoses of asthma and COPD are missing.

Table 4. Inhalation medication in the Netherlands 2020

| *Inhaler type* | *Number of patients ** | *Number of DDDs *** |
|---|---|---|
| Pressured Metered-dose (pMDI) | 831,494 | 228,074,487 |
| Non-propellant (NPI) | 798,598 | 224,615,426 |
| Nebulisers (excluded in further analysis) | 23,479 | 6,931,238 |
| pMDI and/or NPI (included) | 1,388,255 | 452,689,914 |
| pMDI and/or NPI and/or nebulisers (total group) | 1,392,743 | 459,621,151 |

** Users may use different types of inhalers at the same time*

*** Defined daily dose*

In addition 549224 spacers have been issued to 509650 (pMDI using) patients, so 61% of them received a new, yearly-to-be-replaced inhaler.

Table 5. Patient groups not eligible for pMDI to NPI replacement (nebulizers were excluded)

| *Patient group* | *Number of patients* | *Their consumption of pMDI medication (in DDD)* | *Percentage of their pMDI consumption as part of all pMDI consumption* |
|---|---|---|---|
| Severe COPD (COPD and at least 42 DDD prednisone per year) | 45,884 | 27,309,586 | 12.0% |
| Younger than 7 years of age | 74,177 | 5,274,243 | 2.3% |
| All others | 1,392,743 | 195490658 | 85.7% |
| | | 228,074,487 | 100% |

**Outcome of step 3**

Table 6. Number of canisters per group, calculated with product specifications

| *Inhalation medication group* | *Number of pMDI DDDs* | *Number of pMDI canisters* |
|---|---|---|
| ICS | 62,063,901 | 1,194,372 |
| LABA | 15,976,834 | 364,611 |
| LABA-ICS | 71,397,781 | 2,211,663 |
| LABA-LAMA-ICS | 9,256,871 | 308,380 |
| SABA | 48,382,999 | 1,935,320 |
| SABA-ICS | 0 | 0 |
| SAMA | 13,846,309 | 414,560 |
| Total | 220,924,694 | 6,428,906 |

The underlying calculations are at product level, and are not shown.

Table 7. Reduction of $CO_2$ equivalents due to theoretical 70% exchange of pMDI for NPI

| | *Using conversion table from Yeswani* | *Using conversion table from Wilkinson* |
|---|---|---|
| Kilogram CO2 equivalent | 109,942,364 | 119,739,069 |
| 70% reduction of pMDI use (in Kilogram CO2 equivalent) | 76,959,654 | 83,817,348 |

**Outcome of step 4**

Table 8. Financial impact of 70% replacement of pMDI to NPI

| | *DDDs of pMDI, which can be replaced* | *Portion of pMDI to be replaced (70%)* | *Low-cost scenario* | *Average-cost scenario* |
|---|---|---|---|---|
| Volume in DDD | 220,924,694 | 154,647,286 | 154,647,286 | 154,647,286 |
| Medication cost | € 171,372,329 | € 119,960,630 | € 82,861,418 | € 141,025,299 |
| Cost of spacers | € 18,095,091 | € 12,666,564 | 0 | 0 |
| Total cost | € 189,467,420 | € 132,627,194 | € 82,861,418 | € 141,025,299 |
| Impact of exchange | | | € 49,765,776 savings | € 8,398,105 increased costs |

The low-cost scenario would result in € 49.8 million annual savings, the average-cost scenario would result in € 8.4 million annual extra expenditure.
